# Physiological Aging of the Respiratory System (PARS): from development to application

**DOI:** 10.64898/2026.06.15.26355186

**Authors:** Smitha Edakalavan, Jessica Bon, S Mehdi Nouraie

## Abstract

**Background:** Aging has a critical role in lung changes and the outcome of lung disease. Several lung aging equations have been proposed to measure deviation from physiological aging of the respiratory system. In this study, we aimed to develop a single measure of accelerated lung aging and show its application as a measure of lung aging.

**Method:** We used a pre-bronchodilator pulmonary function test (PFT) from NHANES adult participants recruited from 2007 to 2011. We applied Klemera-Dubal Method (KDM) to four PFT measurements, FEV1, FVC, FEF_25-75_, and PEF, to calculate a measure of lung biological aging. Physiological Aging of the Respiratory System (PARS) was calculated from the residual method vs. chronological age. We tested the construct validity of PARS by measuring its association with risk factors of lung health. The prognostic validity was measured using a survival analysis. Sampling weights were applied to all analyses.

**Results:** In 14,123 adult participants, the mean (SD) of accelerated lung age (PARS) was 0 (8.2) years. Participants with a history of asthma and emphysema had 4- and 10-year higher PARS. Cigarette smoking, lower socioeconomic status, black race, higher serum cadmium, and lower serum selenium and magnesium were associated with higher PARS. During 116 months of follow-up, PARS was associated with a higher mortality (HR = 1.06, 95%CI: 1.05-1.07 per year). Females with higher PARS had a higher risk of death (P for interaction < 0.001). Results were consistent across different subgroups and sensitivity analyses.

**Conclusion:** PARS is a noninvasive lung aging marker and can be applied as a single measure of lung accelerated aging in the adult population. Its strong construct and predictive validity support its future application among different populations with and without lung disease.

## Background

Biological age (BA) is a measure of personal vitality and considers various parameters of physiological systems^1^. As a modifiable measure, BA reflects a decline in human bodily function and offers an advantage over chronological age as an intervention outcome and for identifying at-risk populations. In the last few decades, different methodologies have been applied to derive a measure of systemic biological aging. Each measure uses different markers, such as blood chemistry, DNA methylation, and functional measurements^2^.

The respiratory system undergoes various physiological and structural changes with age. Lung function declines with aging after early adulthood. The most important age-related physiological changes are a reduction in static elastic recoil and lung compliance, as well as a reduction in respiratory muscle strength^3-5^. This reduction reflects the normal aging process in the absence of pathology. Aging is an important risk factor for lung diseases, such as Chronic Obstructive Pulmonary Disease (COPD) and interstitial lung disease^6^.

The age-related changes in pulmonary function vary among different individuals and are impacted by several factors, such as genetics, early life events, lifestyle factors, environmental exposures, and diseases^7^. A non-invasive lung BA could assess the variability of age-related changes among individuals and facilitate translational research of the respiratory system. Lung age was first introduced in 1985 as a risk communication tool to help with smoking cessation^8^. Four separate measures of PFT deviation from normal aging trajectories were used to measure the lung age. Application of these four measures increases the variability and poses several limitations for risk stratification and clinical studies. The full range of lung physiology should be covered by measuring the deviation from the normal aging lung. This measure will help to assess the impact of various exposures and preventive strategies on lung health and to risk-stratify individual patients. In this study, we aimed to develop a single measure of lung aging, to establish how it relates to various risk factors of lung disease, and to assess the prognostic ability of this measure.

## Method

We obtained pulmonary function test (PFT) data for adult participants of three NHANES survey cycles (2007, 2009, 2011). Exclusion for the spirometry component of NHANES included chest pain, recent major surgery, or a physical problem with forceful expiration. Spirometry was performed in the standing position using ATS guidelines^9^. The final analytic sample included 14,123 participants. Details of recruitment, procedures, population characteristics, and study design for NHANES are provided through the Centers for Disease Control and Prevention^10^. Mortality data were extracted using the National Death Index from the National Center for Health Statistics (NCHS) data linkage program through December 31, 2022^11^. The NHANES study protocols are approved by the Institutional Review Board of the NCHS, and written informed consent is obtained from participants.

### Statistical analysis

We used pre-bronchodilator FEV1, FVC, FEF_25-75_, and PEF at baseline values. To remove the effect of common genetic and lifestyle factors between height and aging, we used linear regression to adjust each PFT raw value for standing height for each sex. We then built four regression models to predict the chronological age from height-adjusted PFT for each sex. We applied these regressions to the full analysis cohort to comply with NHANES sample weights. The intercept, slope, and root mean square error (RMSE) of each model were used to calculate the lung physiological age with the Klemera-Dubal Method (KDM) ^12, 13^. We then regressed the lung physiological age against the chronological age to calculate the lung accelerated age from the residuals. The accelerated age will be called the Physiological Age of the Respiratory System (PARS) throughout this manuscript. Mean (SEM) and proportion of continuous and categorical variables were presented using the NHANES sampling weight. We used weighted regression to test the association between clinical and demographic factors and PARS. The association between PARS as a continuous variable and time to death was measured using Cox proportional hazard, with appropriate sampling weights. The Cox assumptions, including linearity, were tested. All statistical analyses were performed in Stata 19.5 (StataCorp., College Station, TX).

## Results

We used PFTs of 14,123 NHANES adult (≥18 years) participants. The mean (SEM) age was 44 (0.4), 50% were Male. The proportion of participants with a history of lung disease was 18%, and 40% smoked cigarettes daily (Table 1).

**Table 1.** Characteristics of analytic sample.

| **Table 1- Characteristics of analytic sample.** |  |  |  |
| --- | --- | --- | --- |
|  | Total | Male | Female |
| Age, year* | 44 (0.4) | 43 (0.4) | 45 (0.4) |
| Height, cm* | 169 (0.1) | 176 (0.2) | 163 (0.2) |
| Race/Ethnicity** |  |  |  |
| Hispanic | 14.1 | 15.0 | 13.3 |
| White | 67.6 | 67.6 | 67.6 |
| Black | 11.4 | 10.6 | 12.2 |
| Other | 6.8 | 6.8 | 6.9 |
| Education** |  |  |  |
| Less than high school | 16.8 | 17.7 | 15.9 |
| High school or GED | 22.3 | 23.4 | 21.2 |
| College or AA degree | 31.2 | 29.6 | 32.9 |
| College graduate or above | 29.6 | 29.3 | 30.0 |
| Current Smoking** |  |  |  |
| No | 51.8 | 51.5 | 52.2 |
| Every day | 40.3 | 40.0 | 40.6 |
| Some day | 7.9 | 8.5 | 7.2 |
| Number of days smoked in the last 5 days* | 4.3 (0.04) | 4.3 (0.05) | 4.5 (0.05) |
| Ratio of income to poverty* | 3.0 (0.1) | 3.1 (0.1) | 3.0 (0.1) |
| FEV1, mL* | 3237 (13.6) | 3758 (18.3) | 2722 (13.1) |
| FVC, mL/sec* | 4133 (14.8) | 4848 (19.3) | 3425 (14.7) |
| FEV<sub>25-75</sub>, mL* | 3060 (25.0) | 3445 (31.7) | 2680 (23.6) |
| PEF, mL* | 8294 (37.6) | 9334 (44.9) | 6967 (38.0) |
| History of lung disease** | 17.5 | 16.9 | 18.4 |
| History of asthma** | 14.2 | 12.2 | 16.1 |
| History of emphysema** | 1.2 | 1.5 | 1.1 |
| Death** | 5.9 | 7.2 | 4.6 |
| * Mean (SEM) |  |  |  |
| **Percentage from column. They do not add up to 100 due to rounding. |  |  |  |

The height-adjusted PFT values by subject’s sex are presented in Table S1. The height-adjusted PFTs and age had a strong negative correlation in both males and females, ranging from -0.37 (PEF in males) to - 0.68 (FEV1 in females) (Table S2). These correlations justify the application of KDM to calculate the measure of lung physiological aging.

The average (SD, range) lung age in males and females was 41 (11.9, -10-89) and 59 (8.5, 23-91) (Figure 1A). The correlation between lung age and chronological age was 0.6 in both sexes. The accelerated age (PARS) in the total population ranged from -40 to 49 years (mean 0 and SD = 8.2) (Figure 2).

**Figure 1.**
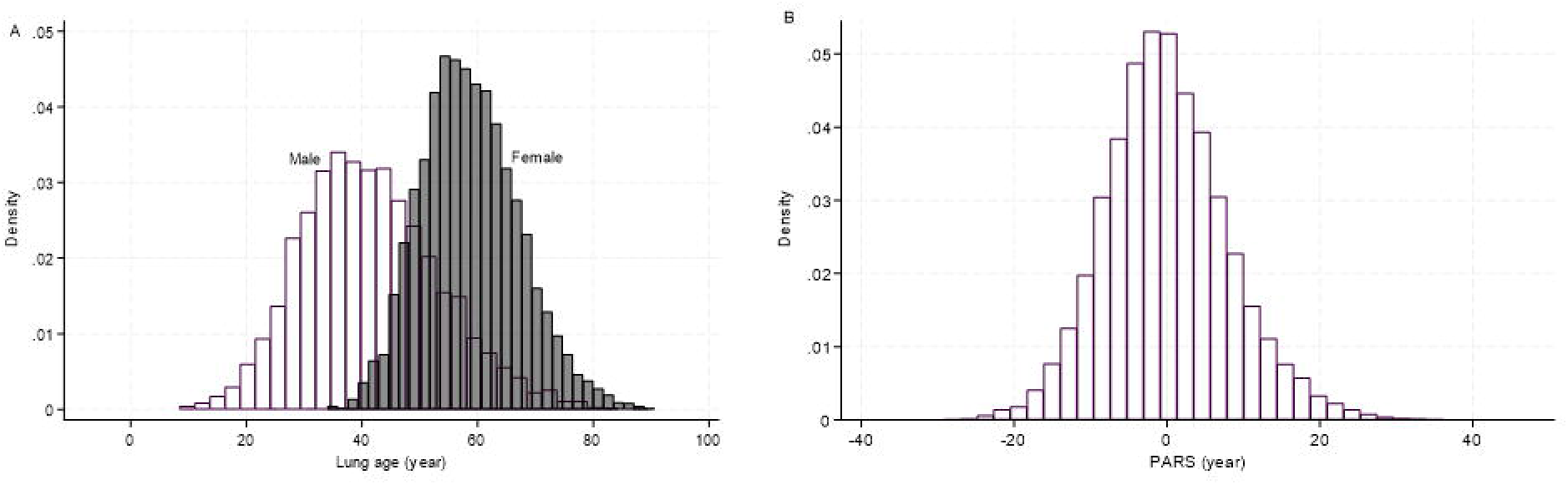
Distribution of A) lung age by sex, B) Accelerated lung age (PARS)

**Figure 2.**
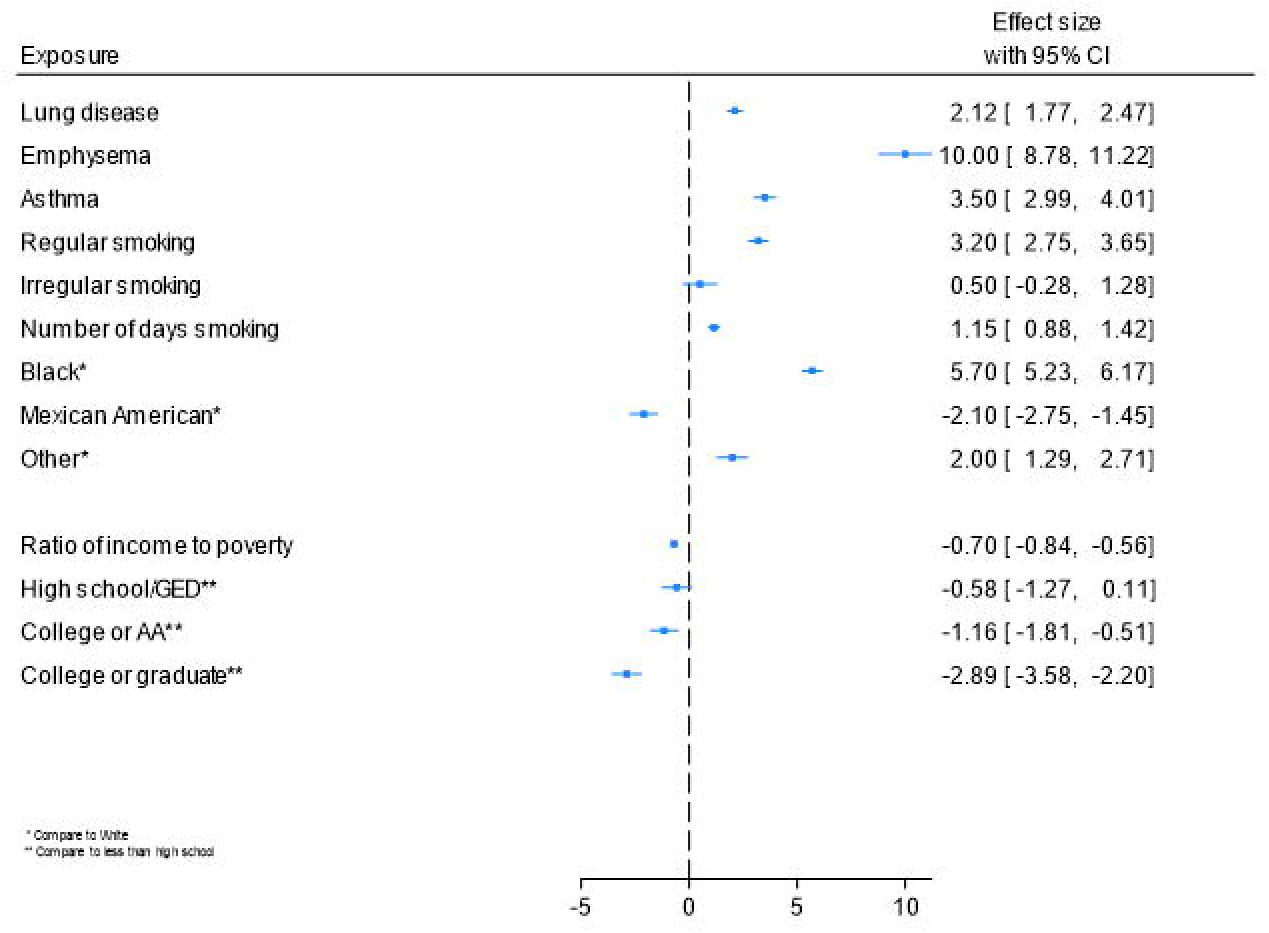
The clinical and socioeconomic predictors of PARS

In participants with a history of lung diseases, the PARS was 2.1 years higher. These values were 10.0 and 3.5 years in emphysema and asthma, respectively. Regular smoking was associated with a 3.1 year higher PARS. Ratio of income to poverty, race, and education was also associated with PARS (Figure 2).

We also analyzed the association between accelerated lung aging and blood metal concentration. Blood cadmium (r = 0.17, N = 13,511) and lead (r = 0.03, N = 13,511) were strongly associated with higher PARS, whereas selenium (r = -0.07, N = 4382), mercury (r = -0.04, N = 13,511), and magnesium (r = -0.07, N = 4382) had a negative association with PARS.

Over a median of 116 (IQR: 99-134) months of follow-up, the risk of death was 5.9% (4.6% in females and 7.2% in males). This risk increases with a positive PARS (HR = 1.06, 95% 1.05-1.07, P<0.001 for each year). We additionally tested for interaction between PARS and participant sex. Accelerated ageing between 0 and 15 years had more effect on males’ death, whereas after 15 years, the women were at a higher risk of death (P for interaction < 0.001, Figure 3). The proportional hazard assumption was violated (P = 0.005), with the effect of PARS on mortality strengthening over time (HR = 1.04 before 134 and 1.09 over 134 months). Adding time interaction did not change the PARS*sex interaction.

**Figure 3.**
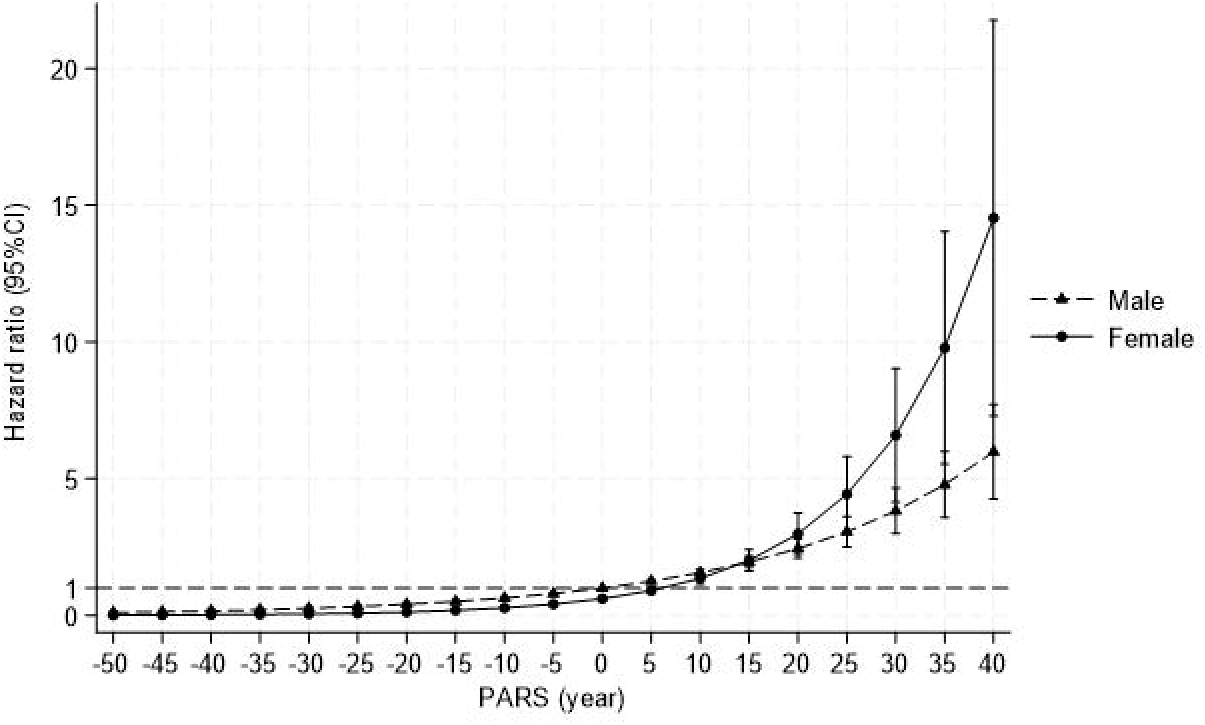
Association between PARS and mortality risk by sex

### Sensitivity and subgroup analysis

We removed the at-risk population to assess the robustness of survival analysis. The association between PARS and higher mortality remains unchanged in non-smokers, participants without any history of respiratory disease, participants with more than a high school education, or Whites (P value for interaction for each subgroup analysis ≥0.05).

In another sensitivity analysis, we only evaluated PFTs with A and B quality ratings (i.e., meet or exceed ATS quality standard, N = 7790). The average PARS in this subgroup was -0.7 (SD = 7.7, ranging from -36 to 49 years). A history of lung diseases (1.7 higher PARS), asthma (3.1 higher PARS), and emphysema (9.3 higher PARS) remained associated with high PARS. Results for SES factors remained unchanged. In this subgroup, the higher PARS remained associated with a higher risk of death (HR = 1.06, P < 0.001). To assess the robustness of the model to the choice of PFT measures, FEF_25-75_ was replaced with FEV6, and the analysis was repeated. In this analysis, the mean (SD) lung age was 41 (11.9) in males and 59 (8.5) in females. This replacement did not impact the results (data are not shown).

## Discussion

In this study, we used four common PFT values to calculate a single lung aging measure and lung age acceleration. The construct validity of lung age acceleration, as represented by PARS, was confirmed by testing the association with known clinical, lifestyle, and environmental risk factors of lung health. We also showed that PARS has a strong prognostic value for death in both participants with and without risk factors of lung disease.

Aging is the single most important risk factor for chronic lung diseases. Genomic instability, telomere attrition, epigenetic alterations, deregulated nutrient sensing, mitochondrial dysfunction, cellular senescence, and stem cell exhaustion are among the important hallmarks of aging in the lung^6^. Among these, the epigenetic clock is shown to be associated with nutrient sensing, mitochondrial activity, and stem cell composition, but not with cellular senescence and telomere attrition^14^.

A biomarker of aging is a noninvasive measure that predicts physiological function in an age-related way^1, 15^. Serum proteins and tissue-specific markers have been used to measure organ-specific aging, including the lungs. The lung tissue methylation clock was highly correlated to chronological age, measured by methylation age^16^. However, lung aging was associated with mortality to a lesser degree compared to other organs, such as the heart and brain^17^. In addition, chronic lung diseases have been associated with other measures of biological age to a lesser degree than what we found in this study. In one study among NHANES participants, the methylation-based biological age was 4 years older in emphysema, and about 1 year older in asthma^18^, much lower than the effect sizes of chronic lung disease on PARS in the current study. These justify the need for a lung-specific measure of aging to be used.

Smoking ^19^ and socioeconomic status (SES) ^20, 21^ are among the most important predictors of biological aging, along with early life factors, such as air pollution and reduced lung growth^22, 23^. Lower SES and air pollution are linked to lower PFT values^24-27^ and chronic lung disease^28^. Diverse forms of stress, ranging from pregnancy, infection^29^, and environmental hazardous exposures^30^, are linked to a rapid increase in the biological age ^31^. Strong associations between smoking and lower SES support that PARS could be used as a single measure of lung aging in this population to study risk factors.

Higher cadmium, lower selenium, and lower magnesium have been shown to affect lung health and reduce PFT measures through several mechanisms. Cadmium increases the oxidative stress in the lung, whereas selenium is a potent antioxidant^32, 33^. Magnesium has several effects on lung health through antioxidant activity, improved mucociliary function, and reduced airway obstruction^34^. In addition, selenium and magnesium have potential anti-aging effects^35, 36^. The main source of blood mercury in humans is shown to be seafoods^37^; therefore, we believe the protective mercury effect on lung aging is derived from nutrition in this study.

Accelerated lung aging is an important prognostic factor in the current study. In the healthy adult population, PFT measures are consistently associated with overall mortality^38-40^. In addition, biological aging mediates the effect of chronological age and mortality in chronic lung diseases^41, 42^. A significant interaction likewise exists between smoking and sex on mortality, with female current smokers showing higher relative risks of all-cause and cardiovascular mortality compared to male current smokers, despite overall survival being higher in females^43^. These findings confirm that the PARS could be used as a unified prognostic measurement of lung health in adults.

This study has several strengths. We derived a single measure of accelerated aging in a large, representative population of healthy adults and demonstrated its robustness to population health status. Unlike other measures of lung age that rely on developing polynomial equations from a limited number of PFT measures in a specific population^8, 44-46^, we calculated PARS from multiple PFT measures, each representing different lung physiology. Therefore, we anticipate that PARS assesses a broader spectrum of lung changes and increases the reliability of measurement. Various PFT measures are highly correlated, which reduces the reliability of regression methods. We used KDM to calculate PARS. KDM is a flexible method of calculating biological aging that applies to different clinical and research settings, and it is not impacted by the collinearity issue. The flexibility of this method was confirmed by alternating between different PFT measures.

There are potential limitations for PARS. Its variance is relatively high compared to some other measures of accelerated aging. PARS does not include measures of pulmonary vascular function and gas exchange. Although these measurements are less frequently available in the healthy population, we plan to incorporate DLco and other measures of vascular function in future iterations.

### Conclusion and future steps

PARS is calculated from lung physiological measures that are age-related. It can be used as a non-invasive marker of accelerated lung aging in the adult population. PARS could be used to study genetic, clinical, and lifestyle predictors of lung function. In addition, it predicts mortality beyond the chronological age. In the future, we plan to test its validity in other populations, including patients with chronic lung disease. After external validation, PARS could be used for clinical and epidemiological studies. Its application for risk stratification, communication, exposomic studies^47^, and clinical trial population enrichment would help to shift the paradigm of lung aging health.

## Data Availability

Data from this project is available from the NHANES website. All statistical codes will be available upon request from the corresponding author.

## Abbreviations

FEV1: Forced Expiratory Volume in one second
FVC: Forced Vital Capacity
FEF_25-75_: Forced Expiratory Flow between 25% and 75%
PEF: Peak Expiratory Flow
FEV6: Forced Expiratory Volume in 6 seconds
DLco: Diffusing Capacity of the Lungs for Carbon Monoxide
NHANES: National Health and Nutrition Examination Survey
PFT: Pulmonary Function Test
ATS: American Thoracic Society
SEM: Standard Error of Mean
SES: Socio-Economic Status

## References

1. Moqri M, Herzog C, Poganik JR, Biomarkers of Aging C, Justice J, Belsky DW, Higgins-Chen A, Moskalev A, Fuellen G, Cohen AA, Bautmans I, Widschwendter M, Ding J, Fleming A, Mannick J, Han JJ, Zhavoronkov A, Barzilai N, Kaeberlein M, Cummings S, Kennedy BK, Ferrucci L, Horvath S, Verdin E, Maier AB, Snyder MP, Sebastiano V, Gladyshev VN. Biomarkers of aging for the identification and evaluation of longevity interventions. Cell. 2023;186(18):3758–75. doi: 10.1016/j.cell.2023.08.003. PubMed PMID: 37657418; PMCID: PMC11088934.

2. Zurbuchen R, von Daniken A, Janka H, von Wolff M, Stute P. Methods for the assessment of biological age - A systematic review. Maturitas. 2025;195:108215. Epub 20250207. doi: 10.1016/j.maturitas.2025.108215. PubMed PMID: 39938306.

3. Janssens JP, Pache JC, Nicod LP. Physiological changes in respiratory function associated with ageing. Eur Respir J. 1999;13(1):197–205. doi: 10.1034/j.1399-3003.1999.13a36.x. PubMed PMID: 10836348.

4. Hankinson JL, Odencrantz JR, Fedan KB. Spirometric reference values from a sample of the general U.S. population. Am J Respir Crit Care Med. 1999;159(1):179–87. doi: 10.1164/ajrccm.159.1.9712108. PubMed PMID: 9872837.

5. Sharma G, Goodwin J. Effect of aging on respiratory system physiology and immunology. Clin Interv Aging. 2006;1(3):253–60. doi: 10.2147/ciia.2006.1.3.253. PubMed PMID: 18046878; PMCID: PMC2695176.

6. Baker JR, Beaulieu D, Avci E, Huang E, Eickelberg O, Meiners S, Savai R, Lehmann M, Konigshoff M. Hallmarks of the ageing lung -10 years later. Eur Respir J. 2026. Epub 20260312. doi: 10.1183/13993003.01272-2025. PubMed PMID: 41819537.

7. International Consortium to Classify Ageing-Related Pathologies respiratory working group m, Weight CM, Tatler AL, Huckstepp RTR, Adcock IM, Barnes PJ, van Beek EJR, Christopher G, Clifford RL, Dhasmana DJ, Hart SP, Hashmat A, Noble S, Sayers I, Short E, Stinson R, Calimport SRG, Bentley BL. The crucial role of normative ageing in respiratory pathogenesis. Lancet Healthy Longev. 2026;7(4):100842. Epub 20260424. doi: 10.1016/j.lanhl.2026.100842. PubMed PMID: 42044655.

8. Morris JF, Temple W. Spirometric “lung age” estimation for motivating smoking cessation. Prev Med. 1985;14(5):655–62. doi: 10.1016/0091-7435(85)90085-4. PubMed PMID: 4070195.

9. Miller MR, Hankinson J, Brusasco V, Burgos F, Casaburi R, Coates A, Crapo R, Enright P, van der Grinten CP, Gustafsson P, Jensen R, Johnson DC, MacIntyre N, McKay R, Navajas D, Pedersen OF, Pellegrino R, Viegi G, Wanger J, Force AET. Standardisation of spirometry. Eur Respir J. 2005;26(2):319–38. doi: 10.1183/09031936.05.00034805. PubMed PMID: 16055882.

10. Centers for Disease Control and Prevention (CDC). National Center for Health Statistics (NCHS). National Health and Nutrition Examination Survey Data: Hyattsville, MD: U.S. Department of Health and Human Services, Centers for Disease Control and Prevention. ; 2007–2011 [6/2/2026]. Available from: https://www.cdc.gov/nchs/nhanes/index.htm.

11. Centers for Disease Control and Prevention. Public-use linked mortality files. 2026 [6/2/2026]. Available from: https://ftp.cdc.gov/pub/Health_Statistics/NCHS/datalinkage/linked_mortality/.

12. Klemera P, Doubal S. A new approach to the concept and computation of biological age. Mech Ageing Dev. 2006;127(3):240–8. Epub 20051128. doi: 10.1016/j.mad.2005.10.004. PubMed PMID: 16318865.

13. Kwon D, Belsky DW. A toolkit for quantification of biological age from blood chemistry and organ function test data: BioAge. Geroscience. 2021;43(6):2795–808. Epub 20211102. doi: 10.1007/s11357-021-00480-5. PubMed PMID: 34725754; PMCID: PMC8602613.

14. Kabacik S, Lowe D, Fransen L, Leonard M, Ang SL, Whiteman C, Corsi S, Cohen H, Felton S, Bali R, Horvath S, Raj K. The relationship between epigenetic age and the hallmarks of aging in human cells. Nat Aging. 2022;2(6):484–93. Epub 20220516. doi: 10.1038/s43587-022-00220-0. PubMed PMID: 37034474; PMCID: PMC10077971.

15. Nakamura E, Lane MA, Roth GS, Ingram DK. A strategy for identifying biomarkers of aging: further evaluation of hematology and blood chemistry data from a calorie restriction study in rhesus monkeys. Exp Gerontol. 1998;33(5):421–43. doi: 10.1016/s0531-5565(97)00134-4. PubMed PMID: 9762521.

16. Horvath S. DNA methylation age of human tissues and cell types. Genome Biol. 2013;14(10):R115. doi: 10.1186/gb-2013-14-10-r115. PubMed PMID: 24138928; PMCID: PMC4015143.

17. Oh HS, Rutledge J, Nachun D, Palovics R, Abiose O, Moran-Losada P, Channappa D, Urey DY, Kim K, Sung YJ, Wang L, Timsina J, Western D, Liu M, Kohlfeld P, Budde J, Wilson EN, Guen Y, Maurer TM, Haney M, Yang AC, He Z, Greicius MD, Andreasson KI, Sathyan S, Weiss EF, Milman S, Barzilai N, Cruchaga C, Wagner AD, Mormino E, Lehallier B, Henderson VW, Longo FM, Montgomery SB, Wyss-Coray T. Organ aging signatures in the plasma proteome track health and disease. Nature. 2023;624(7990):164–72. Epub 20231206. doi: 10.1038/s41586-023-06802-1. PubMed PMID: 38057571; PMCID: PMC10700136.

18. Perez-Garcia J, Khodasevich D, De Matteis S, Rice MB, Needham BL, Rehkopf DH, Cardenas A. Epigenetic age among US adults with chronic respiratory diseases: results from NHANES 1999-2002. Eur Respir J. 2025;65(6). Epub 20250619. doi: 10.1183/13993003.00293-2025. PubMed PMID: 40441879; PMCID: PMC12177333.

19. Mamoshina P, Kochetov K, Cortese F, Kovalchuk A, Aliper A, Lane E, Scheibye-Knudsen M, Cantor CR, Skjodt NM, Kovalchuk O, Zhavoronkov A. Blood Biochemistry Analysis to Detect Smoking Status and Quantify Accelerated Aging in Smokers. Sci Rep. 2019;9(1):142. Epub 20190115. doi: 10.1038/s41598-018-35704-w. PubMed PMID: 30644411; PMCID: PMC6333803.

20. Dalecka A, Bartoskova Polcrova A, Pikhart H, Bobak M, Ksinan AJ. Living in poverty and accelerated biological aging: evidence from population-representative sample of U.S. adults. BMC Public Health. 2024;24(1):458. Epub 20240213. doi: 10.1186/s12889-024-17960-w. PubMed PMID: 38350911; PMCID: PMC10865704.

21. Pala D, Xie Y, Xu J, Shen L. Modeling the impact of socioeconomic disparity, biological markers and environmental exposures on phenotypic age using mediation analysis and structural equation models. Int J Med Inform. 2025;193:105661. Epub 20241028. doi: 10.1016/j.ijmedinf.2024.105661. PubMed PMID: 39481175; PMCID: PMC11973974.

22. Eminson K, Chen Y, Granell R, Gulliver J, Cai YS, Hansell AL. Prenatal and early life exposure to air pollution and lung function development aged 8-24 years: The ALSPAC study. Environ Res. 2026;299:124278. Epub 20260317. doi: 10.1016/j.envres.2026.124278. PubMed PMID: 41850474.

23. Hu CY, Alcala CS, Lamadrid-Figueroa H, Mercado-Garcia A, Tamayo-Ortiz M, Gutierrez-Avila I, Kloog I, Just AC, He MZ, Yitshak-Sade M, Rivera-Rivera NY, Estrada-Gutierrez G, Tellez-Rojo MM, Wright RO, Wright RJ, Rosa MJ. Identifying Critical Windows and Joint Effects of Prenatal Air Pollution and Temperature Exposure and Lung Function in Schoolchildren: Findings From a Prospective Birth Cohort Study. Chest. 2026;169(1):179–93. Epub 20250910. doi: 10.1016/j.chest.2025.08.022. PubMed PMID: 40939936; PMCID: PMC12809656.

24. Xue S, Broerman MJ, Goobie GC, Kass DJ, Fabisiak JP, Wenzel SE, Nouraie SM. Gaseous Air Pollutants and Lung Function in Fibrotic Interstitial Lung Disease (fILD): Evaluation of Different Spatial Analysis Approaches. Environ Sci Technol. 2025;59(12):5936–45. Epub 20250322. doi: 10.1021/acs.est.4c11275. PubMed PMID: 40119855; PMCID: PMC11966764.

25. Rosser FJ, Han YY, Forno E, Guilbert TW, Bacharier LB, Phipatanakul W, Goobie GC, Nouraie SM, Martinez M, Celedon JC. Long-Term Exposure to Particulate Matter <2.5 mum and Lung Function Change in Children with Asthma Receiving Inhaled Corticosteroids. Am J Respir Crit Care Med. 2023;208(5):622–4. doi: 10.1164/rccm.202303-0371LE. PubMed PMID: 37311241; PMCID: PMC12042981.

26. Goobie GC, Ryerson CJ, Johannson KA, Schikowski E, Zou RH, Khalil N, Marcoux V, Assayag D, Manganas H, Fisher JH, Kolb MRJ, Gibson KF, Kass DJ, Zhang Y, Lindell KO, Nouraie SM. Neighborhood-Level Disadvantage Impacts on Patients with Fibrotic Interstitial Lung Disease. Am J Respir Crit Care Med. 2022;205(4):459–67. doi: 10.1164/rccm.202109-2065OC. PubMed PMID: 34818133.

27. Goobie GC, Carlsten C, Johannson KA, Khalil N, Marcoux V, Assayag D, Manganas H, Fisher JH, Kolb MRJ, Lindell KO, Fabisiak JP, Chen X, Gibson KF, Zhang Y, Kass DJ, Ryerson CJ, Nouraie SM. Association of Particulate Matter Exposure With Lung Function and Mortality Among Patients With Fibrotic Interstitial Lung Disease. JAMA Intern Med. 2022;182(12):1248–59. doi: 10.1001/jamainternmed.2022.4696. PubMed PMID: 36251286; PMCID: PMC9577882.

28. Park SL, Lichtensztajn D, Yang J, Wu J, Shariff-Marco S, Stram DO, Inamdar P, Fruin S, Larson T, Tseng C, Setiawan VW, Gomez SL, Samet J, Le Marchand L, Wilkens LR, Ritz B, Wu AH, Cheng I. Ambient Air Pollution and Chronic Obstructive Pulmonary Disease: The Multiethnic Cohort Study. Ann Am Thorac Soc. 2025;22(5):698–706. doi: 10.1513/AnnalsATS.202404-387OC. PubMed PMID: 39847697; PMCID: PMC12051916.

29. Horvath S, Levine AJ. HIV-1 Infection Accelerates Age According to the Epigenetic Clock. J Infect Dis. 2015;212(10):1563–73. doi: 10.1093/infdis/jiv277. PubMed PMID: 25969563; PMCID: PMC4621253.

30. Gross A, Tham R, Dharmage SC, Roosli M, Frey U, Gorlanova O. Exposure to long-term ambient air pollution and lung function in adults: a systematic review and meta-analysis. Eur Respir Rev. 2025;34(176). Epub 20250611. doi: 10.1183/16000617.0264-2024. PubMed PMID: 40500128; PMCID: PMC12152583.

31. Poganik JR, Zhang B, Baht GS, Tyshkovskiy A, Deik A, Kerepesi C, Yim SH, Lu AT, Haghani A, Gong T, Hedman AM, Andolf E, Pershagen G, Almqvist C, Clish CB, Horvath S, White JP, Gladyshev VN. Biological age is increased by stress and restored upon recovery. Cell Metab. 2023;35(5):807–20 e5. Epub 20230421. doi: 10.1016/j.cmet.2023.03.015. PubMed PMID: 37086720; PMCID: PMC11055493.

32. Kwon OB, Lee EJ, Lim MN, Kim J, Kim WJ. Relationship between Serum Cadmium Concentration and Lung Function: A Study Using Korea National Health and Nutrition Examination Survey Data. Tuberc Respir Dis (Seoul). 2025;88(4):696–707. Epub 20250731. doi: 10.4046/trd.2024.0161. PubMed PMID: 40744453; PMCID: PMC12488350.

33. Fu YX, Wang YB, Bu QW, Guo MY. Selenium Deficiency Caused Fibrosis as an Oxidative Stress-induced Inflammatory Injury in the Lungs of Mice. Biol Trace Elem Res. 2023;201(3):1286–300. Epub 20220409. doi: 10.1007/s12011-022-03222-6. PubMed PMID: 35397105.

34. Landon RA, Young EA. Role of magnesium in regulation of lung function. J Am Diet Assoc. 1993;93(6):674–7. doi: 10.1016/0002-8223(93)91675-g. PubMed PMID: 8509592.

35. Dominguez LJ, Veronese N, Barbagallo M. Magnesium and the Hallmarks of Aging. Nutrients. 2024;16(4). Epub 20240209. doi: 10.3390/nu16040496. PubMed PMID: 38398820; PMCID: PMC10892939.

36. Bjorklund G, Shanaida M, Lysiuk R, Antonyak H, Klishch I, Shanaida V, Peana M. Selenium: An Antioxidant with a Critical Role in Anti-Aging. Molecules. 2022;27(19). Epub 20221005. doi: 10.3390/molecules27196613. PubMed PMID: 36235150; PMCID: PMC9570904.

37. Chakif D, Furrer J. Heavy Metal Toxicity in Clinical and Environmental Health: Sources, Mechanisms, Diagnostics, and Evidence-Based Management of Mercury, Lead, Cadmium, and Arsenic. Int J Mol Sci. 2026;27(8). Epub 20260414. doi: 10.3390/ijms27083513. PubMed PMID: 42074156; PMCID: PMC13116634.

38. Collaro AJ, Chang AB, Marchant JM, Chatfield MD, Dent A, Blake T, Mawn P, Fong K, McElrea MS. Associations between lung function and future cardiovascular morbidity and overall mortality in a predominantly First Nations population: a cohort study. Lancet Reg Health West Pac. 2021;13:100188. Epub 20210705. doi: 10.1016/j.lanwpc.2021.100188. PubMed PMID: 34527981; PMCID: PMC8403916.

39. Sabia S, Shipley M, Elbaz A, Marmot M, Kivimaki M, Kauffmann F, Singh-Manoux A. Why does lung function predict mortality? Results from the Whitehall II Cohort Study. Am J Epidemiol. 2010;172(12):1415–23. Epub 20101020. doi: 10.1093/aje/kwq294. PubMed PMID: 20961971; PMCID: PMC2998200.

40. Weinmayr G, Schulz H, Klenk J, Denkinger M, Duran-Tauleria E, Koenig W, Dallmeier D, Rothenbacher D, Acti FESG. Association of lung function with overall mortality is independent of inflammatory, cardiac, and functional biomarkers in older adults: the ActiFE-study. Sci Rep. 2020;10(1):11862. Epub 20200717. doi: 10.1038/s41598-020-68372-w. PubMed PMID: 32681112; PMCID: PMC7367870.

41. Pugashetti JV, Kim JS, Bose S, Adegunsoye A, Linderholm AL, Chen CH, Strek ME, Flaherty KR, Murray S, Newton CA, Alqalyoobi S, Ma SF, Mychaleckyj JC, Bowler RP, Han MK, Curtis JL, Martinez FJ, Smith JA, Noth I, Oldham JM. Biological Age, Chronological Age, and Survival in Pulmonary Fibrosis: A Causal Mediation Analysis. Am J Respir Crit Care Med. 2024;210(5):639–47. doi: 10.1164/rccm.202310-1887OC. PubMed PMID: 38843133; PMCID: PMC11389564.

42. Goobie GC, Marinescu DC, Adegunsoye A, Bourbeau J, Carlsten C, Clifford RL, Doiron D, Duan Q, Gibson KF, Grant-Orser A, Hernandez Cordero AI, Johannson KA, Kass DJ, Kim SE, Leung JM, Li X, Tan W, Xi Yang C, Nouraie SM, Ryerson CJ, Hackett TL, Zhang Y. Accelerated epigenetic aging worsens survival and mediates environmental stressors in fibrotic interstitial lung disease. Eur Respir J. 2025. Epub 20250130. doi: 10.1183/13993003.01618-2024. PubMed PMID: 39884761.

43. Vallee A. Associations between tobacco smoking and mortality: a sex-stratified cohort analysis. Eur J Public Health. 2025;35(6):1212–8. doi: 10.1093/eurpub/ckaf194. PubMed PMID: 41134686; PMCID: PMC12707473.

44. Ishida Y, Ichikawa YE, Fukakusa M, Kawatsu A, Masuda K. Novel equations better predict lung age: a retrospective analysis using two cohorts of participants with medical check-up examinations in Japan. NPJ Prim Care Respir Med. 2015;25:15011. Epub 20150319. doi: 10.1038/npjpcrm.2015.11. PubMed PMID: 25789796; PMCID: PMC4373493.

45. Liang X, Xie Y, Gao Y, Zhou Y, Jian W, Jiang M, Wang H, Zheng J. Estimation of lung age via a spline method and its application in chronic respiratory diseases. NPJ Prim Care Respir Med. 2022;32(1):36. Epub 20220929. doi: 10.1038/s41533-022-00293-9. PubMed PMID: 36175436; PMCID: PMC9522795.

46. Khelifa MB, Salem HB, Sfaxi R, Chatti S, Rouatbi S, Saad HB. “Spirometric” lung age reference equations: A narrative review. Respir Physiol Neurobiol. 2018;247:31–42. Epub 20170901. doi: 10.1016/j.resp.2017.08.018. PubMed PMID: 28870870.

47. Chung MK, House JS, Akhtari FS, Makris KC, Langston MA, Islam KT, Holmes P, Chadeau-Hyam M, Smirnov AI, D. X, Thessen AE, Cui Y, Zhang K, Manrai AK, Motsinger-Reif A, Patel CJ, Members of the Exposomics C. Decoding the exposome: data science methodologies and implications in exposome-wide association studies (ExWASs). Exposome. 2024;4(1):osae001. Epub 20240117. doi: 10.1093/exposome/osae001. PubMed PMID: 38344436; PMCID: PMC10857773.

