## Supplementary Tables 1 and 2 for "Physiological Aging of the Respiratory System (PARS): from development to application"

| **Table S1- Mean (SEM) of height-adjusted PFT values** | | |
| --- | --- | --- |
|  | Male | Female |
| FEV1, mL* | 3740 (16.5) | 2701 (13.0) |
| FVC, mL/sec* | 4823 (14.8) | 3398 (12.7) |
| FEV_25-75_, mL* | 3433 (32.0) | 2666 (24.8) |
| PEF, mL* | 9601 (42.7) | 6931 (37.1) |

| **Table S2- The correlation coefficient between height-adjusted PFT values and age (N = 14,123)** | | |
| --- | --- | --- |
|  | Male | Female |
| FEV1 | -0.65 | -0.68 |
| FVC | -0.50 | -0.56 |
| FEV_25-75_ | -0.62 | -0.60 |
| PEF | -0.37 | -0.42 |
